# Type 2 diabetes worsens the outcome of ischemia/reperfusion in female STEMI patients and female db/db mice with HFpEF cardiometabolic phenotype

**DOI:** 10.1101/2025.04.14.25325801

**Authors:** Ivana Iveljic, Megan Young, Elvira Corhodzic, Fenn Cullen, Hiran A. Prag, Michael P. Murphy, Dunja Aksentijevic

**Affiliations:** Faculty of Medicine University of Tuzla, Tuzla, Bosnia and Herzegovina; University Clinical Centre Tuzla, Tuzla, Bosnia and Herzegovina; William Harvey Research Institute, Barts and the London Faculty of Medicine and Dentistry, Queen Mary University of London; London, United Kingdom; MRC Mitochondrial Biology Unit, University of Cambridge, Cambridge, United Kingdom

**Keywords:** type 2 diabetes, STEMI, female patients, db/db mice, HFpEF, metabolism, ischemia/reperfusion

## Abstract

**Background:** Heart failure with preserved ejection fraction (HFpEF) poses a significant global health challenge, disproportionately affecting women. Diabetic women with HFpEF represent a high-risk subgroup, particularly after experiencing ST-segment elevation myocardial infarction (STEMI), exhibiting increased mortality compared to men. While prolonged door-to-balloon (DTB) times, reflecting delayed reperfusion, are a critical factor in STEMI outcomes, they alone do not fully capture the observed outcome variability in diabetic women. Using an integrated clinical and pre-clinical approach this study aimed to investigate the relative contributions of metabolic dysfunction and coronary artery disease (CAD) in type 2 diabetes (T2D) to STEMI outcomes in women, beyond the impact of DTB time.

**Methods:** A retrospective case-control study analysed female STEMI patients undergoing primary percutaneous coronary intervention (pPCI, n=40 T2D, n=40 non-diabetic controls), comparing clinical characteristics, treatment strategies, and early outcomes. A preclinical model (female db/db mice) assessed cardiac function via echocardiography, Langendorff perfusions, and ischemia-reperfusion protocols. Metabolome of heart, liver, and skeletal muscle was assessed by ¹H NMR spectroscopy

**Results:** Our study reveals significantly higher mortality, impaired left ventricular function post-PCI, and increased implantable cardioverter-defibrillator (ICD) implantation rates in diabetic STEMI patients, irrespective of DTB time, when compared to non-diabetic controls. Elevated inflammatory markers, acute hyperglycaemia and evidence of cardio-hepatic damage were identified in T2D patients. db/db mice exhibited analogous T2D-associated pathophysiology, including increased ischemia-reperfusion injury exacerbated by metabolic disturbances in the myocardium, liver, and skeletal muscle versus non-diabetic controls

**Conclusions:** In diabetic women, multiple factors beyond reperfusion delays exacerbate acute myocardial injury. This necessitates the development of sex-specific strategies to manage the cardiovascular complications of diabetic HFpEF. The db/db mouse model provides a relevant preclinical tool for future research as mimics human T2D-associated HFpEF and STEMI outcome.

## Background

Heart failure with preserved ejection fraction (HFpEF) poses a significant and growing global health burden with increasing prevalence, largely due to aging populations as well as the rising incidence of contributing factors including obesity, hypertension and type 2 diabetes (T2D)^1,2^. Women make up the majority of the HFpEF population, exhibiting relatively high burden of morbidity and mortality^3^. Phenotypic profile of HFpEF in women is defined by comorbidities, including older age, obesity, atrial fibrillation, inflammation, hypertension, hyperlipidaemia and T2D^3–6^. In women, T2D has been shown to elevate the risk of developing HFpEF by five times, compared to a 2.4-fold increase in men^7^. Furthermore, female sex is independently associated with the presence of diastolic dysfunction and worse clinical outcomes in HFpEF, highlighting that a sex-specific approach is key to investigating the pathophysiology of HFpEF.

The intersection of coronary artery disease (CAD) and metabolic disorders in T2D increases the risk of acute coronary syndrome (ACS), but also significantly aggravates the course of myocardial infarction (MI) worsening patient prognosis^8^. Among the affected populations, female patients with T2D represent a particularly vulnerable group, exhibiting disproportionately poorer outcomes including increased mortality after ST-segment elevation myocardial infarction (STEMI)^8–13^. This heightened risk in women is potentially attributed to the cumulative burden of metabolic dysfunction, inflammation, oxidative stress and cardiac remodeling associated with T2D. However, in addition to biological factors, disparities in outcomes may also be due to poorer cardiovascular risk factor management in routine T2D care of women^14^. Although female T2D patients make up a significant proportion of patients with STEMI, clinical studies relatively rarely address differences in the STEMI outcomes in female patients with T2D^15^.

Primary percutaneous coronary intervention (PCI) is the gold standard for achieving rapid reperfusion post-STEMI. The critical nature of prompt intervention is underscored by the established relationship between Door-to-Balloon (DTB) reperfusion times and patient outcomes, where delays have been consistently associated with increased morbidity and mortality^16^. Generally, diabetic women experience delays in recognising and seeking treatment for STEMI, as they may present atypically with fatigue, nausea/vomiting, shortness of breath, or abdominal discomfort, rather than classic chest pain. Prolonged DTB times have been linked to higher mortality rates, regardless of how the patient presents^16^.

Given the heterogeneous pathogenic mechanisms underlying HFpEF in diabetic women, it is unclear whether time to reperfusion (DTB) is the sole determinant of the poor STEMI outcome as in other patient populations. In this study we used an integrated clinical and pre-clinical (STEMI diabetic patients, db/db mouse model of T2D) approach to investigate whether the higher burden of co-morbidities interact synergistically with duration of the ischemic injury to exacerbate myocardial damage and impair function during reperfusion of female diabetic myocardium.

## Methods

### Study population

Population-based retrospective case-control study was conducted at the Department of Interventional Cardiology, Clinic for Invasive Cardiology, University Clinical Centre Tuzla, Tuzla, Bosnia and Herzegovina. The study protocol was approved by the Ethics Committee of University Clinical Centre Tuzla (Ref. 02-09/2-166-2/24) and all methods were performed in accordance with their guidelines and regulations.

Records of 1100 patients who underwent pPCI due to STEMI (ESC 2023 diagnosis criteria)^12^ were screened for inclusion in the study. Demographic data, medical history, laboratory data, and clinical data during hospitalization were retrieved from hospital patient database. Patients with diabetes were identified as those with known (treated or not) T2D prior to admission. Our study involved 80 female patients (mean age 65) with STEMI who underwent cardiac catheterisation with pPCI between January 2019 and December 2023.

We compared baseline clinical characteristics, treatment strategy and outcomes among women with diabetes (n=40, mean age 65 years), with the control arm consisting of age-matched female STEMI non-diabetic patients (n=40). A special focus on the impact of the treatment strategy on early mortality and cardiac function. The clinical characteristics of the patient cohort is shown in Tables 1-5. Excluded were patients with previous MI, pre-diabetes, heart failure history, angioplasty in the last 3 months, heart failure or cardiogenic shock, severe renal failure (serum creatinine >200 mmol/L, eGFR <44 ml/min/1.73 m^2^), cancer and infectious diseases.

**Table 1.** Patient characteristics at the time of hospitalization. Data are presented as mean ± SEM. Normality of data distribution was examined using Shapiro– Wilk’s normality test. Comparison between two groups was performed by Student’s t-test (Gaussian data distribution) or Mann-Whitney U test when data was non-normally distributed. Contingency analysis by chi-squared and Fisher’s exact test (two-sided). *p<0.05 vs control. Abbreviations: OAD-oral antidiabetic therapies including metformin, SGLT-sodium glucose co-transporter.

|  | Control<br>(n=40) | Diabetic<br>(n=40) | P value |
| --- | --- | --- | --- |
| Sex | female | female |  |
| Age (years) | 65±1.3 | 65±1.3 |  |
| BMI (kg/m<sup>2</sup>) | 29.3±0.4 | **\*33±0.5** | 0.0001 |
| HbA1c (%) | 5.4±0.04 | **\*8.78±0.3** | 0.0001 |
| Hypertension | 34 (85%) | 36 (90%) |  |
| Obesity | 12 (15%) | **\*29 (36%)** | 0.0003 |
| CKD | 3 (7.5%) | 9 (22%) |  |
| Stage1 | 1 | 3 |  |
| Stage 2 | 1 | 3 |  |
| Stage 3 | 1 | 3 |  |
| OAD (non-SGLT2) | - | 36 |  |
| SGLT2i | - | 4 |  |
| Insulin | - | 24 |  |

### Coronary Angiography and pPCI Procedure

The pPCI procedure was performed with the adoption of a standard radial approach that uses a 6F-guiding catheter. After bolus of heparin (100 U/Kg), 300 mg aspirin, and P2Y12 inhibitor in loading dose were given (clopidogrel 600 mg or prasugrel 60 mg), target lesion was crossed through 0.014-inch wires. Non-ionic, low-osmolality contrast agent was used in all patients. After angioplasty, all patients were admitted to the Department of Interventional Cardiology. In addition, they were given 100 mg aspirin and 75 mg clopidogrel or 10 mg prasugrel.

Concomitant medical treatment with β-blockers, angiotensin-converting enzyme inhibitors, and statins, were prescribed in accordance with the ESC Guideline for the management of ACS^17^. Door-to-balloon time was defined from patient anamnestic data (duration of the chest pain before entering in the catheterisation laboratory). Echocardiography was performed 4 days post-hospitalization just before discharge, or, in haemodynamically unstable patients, after the index procedure.

Antecubital venous blood samples for the laboratory analysis were collected at the beginning of the angiography procedure in the catheterisation laboratory, and on the third day of the hospitalization. Cardiovascular mortality in patients was defined as death related to myocardial ischemia and infarction, cardiogenic shock, heart failure, arrhythmia (ventricular fibrillation, ventricular tachycardia, or asystole), and cardiac arrest due to other or undetermined causes or cerebrovascular accident.

### Animals

For all experiments in this study, 12-week female db/db mice (n=12) were purchased from Charles River Laboratories (Italy) and aged until 20-weeks. 20-week lean (C57/Bl6) controls (n=12) were purchased from Charles River Laboratories (UK). Animals were housed in individually ventilated cages and maintained under controlled temperature (22 ± 2 °C) on a 12:12 h light and dark cycles with access to standard chow diet (irradiated PicoLab® Mouse Diet 20 EXT, 5R58) and water *ad libitum.* Enrichment was provided in the form of tunnels and chew sticks. All animal experiments were in keeping with the UK Home Office Animals (Scientific Procedures) Act, 1986. At the end of the experimental model induction protocol, all animals were humanely sacrificed by phenobarbital I.P. injection.

### Biochemical analysis

Tissues (liver, skeletal muscle, heart) were rapidly collected upon termination and snap frozen in liquid N_2_ using Wollenberger tongs^18^. The frozen heart weight was normalised to tibia length to assess the degree of cardiac hypertrophy (heart weight:tibia length). Blood samples taken from the thoracic cavity at the time of sacrifice were centrifuged at 3000 rpm to obtain plasma. All samples were stored at –80°C until biochemical analysis. Plasma samples were analysed by the Clinical Biochemistry Laboratory at Addenbrookes Hospital, Cambridge.

### In vivo Assessment of left ventricular diastolic and systolic function

M-mode and Doppler echocardiography was performed in 20-week db/db and lean control mice. Anaesthesia was induced with 4% isoflurane and maintained at 1.5-2% for the duration of the procedure. Before assessment of cardiac function, fur was removed from the chest area to allow accurate assessment of cardiac function and mice were allowed to stabilize for at least 10 minutes. Body temperature was maintained at 37°C. During echocardiography, heart rate was measured from electrocardiogram. Echocardiography images were recorded using a Vevo-3100 imaging system with a 40-MHz linear probe (VisualSonics, Toronto, Canada).

Morphological measurements were taken in the parasternal short axis view at the level of the papillary muscles and ejection fraction was calculated from M-mode images. Diastolic transmitral left ventricle (LV) inflow images were acquired from apical four-chamber views using pulsed-wave doppler to calculate early (E) and late (atrial, A) peak filling blood flow velocities and E-wave deceleration time. Tissue doppler imaging in apical four-chamber view at the mitral annulus allowed measurements of early (e’) and atrial (a’) peak tissue velocities. The E/e’ ratio could then be calculated. Analysis was performed using VevoLab 5.5.1 software.

### Langendorff heart perfusions

Mice were administered terminal anaesthesia via intra-peritoneal pentobarbitone injection. Beating hearts were rapidly excised, cannulated, and perfused in isovolumic Langendorff mode at 80 mmHg pressure maintained by a St. Thomas Hospital peristaltic pump controller feedback system (AD Instruments, UK), with Krebs–Henseleit (KH) buffer continuously gassed with 95% O_2_/5% CO_2_ (pH 7.4, 37°C) containing (in mM): NaCl (116), KCl (4.7), MgSO_4_.7H_2_O (1.2), KH_2_PO_4_ (1.2), NaHCO_3_ (25), CaCl_2_ (1.4), and glucose (11)^19^. Cardiac function was assessed using a fluid-filled cling-film balloon inserted into the LV connected via a line to a pressure transducer and a Powerlab system (AD Instruments). The volume of the intraventricular balloon was adjusted using a 1.0 mL syringe to achieve an initial LV diastolic pressure of 4–9 mmHg. Functional parameters were recorded using LabChart software v.7 (AD Instruments) throughout the experiment. LV developed pressure (LVDP) was calculated from the difference between systolic and diastolic pressure^18–27^.

### Ischemia Reperfusion Protocol

After 20 min of equilibration, Langendorff crystalloid KH buffer perfused hearts were subject to 20 min of global normothermic (37°C) ischemia and 2 hours of reperfusion^18,19,22,24–27^. At the end of the protocol, hearts were perfused for 1 min with 1% triphenyltetrazolium chloride (TTC) in PBS followed by 10 min incubation at 37°C in 1% TTC. Hearts were sectioned into 1 mm slices (mouse heart gauge, Zivic instruments, USA), imaged and infarct size was quantified using Fiji-ImageJ Software ^18,25^.

### Metabolomic analysis using ^1^H Nuclear Magnetic Resonance (NMR) Spectroscopy

Snap frozen and pulverized tissue [liver, skeletal muscle (gastrocnemius and soleus), heart] was analysed using ^1^H NMR high resolution spectroscopy as previously described^18,28^. Pulverised tissue samples (∼45 mg) were subject to methanol/water/chloroform phase extraction. ^1^H NMR spectra were acquired using a vertical-bore, ultra-shielded Bruker 14.1. Tesla (600 MHz) spectrometer with a BBO probe at 298K using the Bruker noesygppr1d pulse sequence. Acquisition parameters were 128 scans, 4 dummy scans and 20.8 ppm sweep width, acquisition time of 2.6s, pre-scan delay of 4s, 90° flip angle and experiment duration of 14.4 minutes per sample. TopSpin (version 4.0.5) software was used for data acquisition and for metabolite quantification. FIDs were multiplied by a line broadening factor of 0.3 Hz and Fourier-transformed, phase and automatic baseline-correction were applied. Chemical shifts were referenced to the TSP signal. Metabolite peaks of interest were initially integrated automatically using a pre-written integration region text file and then manually adjusted where required. Assignment of metabolites to their respective peaks was carried out based on previously obtained in-house data, confirmed by chemical shift and using Chenomx NMR Profiler Version 8.1 (Chenomx, Canada). Peak areas were normalized to the total metabolite peak area. Quantification of glycogen was performed by ^1^H magnetic resonance, giving a measure of the concentration of glucose monomers that are present in the observed peak. Being a large macromolecule, with possible differences in the mobility of glycosyl units, glycogen has been reported to be fully visible by MRS. The modified dual-phase Folch extraction method used for separating aqueous and lipid metabolites was not optimized for the extraction of glycogen, however all samples underwent the same extraction procedure allowing between treatment group comparison^18^.

### Statistical Analysis

Data are presented as mean ± SEM unless otherwise stated. Normality of data distribution was examined using Shapiro–Wilk’s normality test. Comparison between two groups was performed by Student’s t-test (Gaussian data distribution) or Mann-Whitney U test when data was non-normally distributed, and an unequal variance t-test was performed in the absence of equal variance between groups. Two-way analysis of variance (ANOVA) with Tukey’s correction for multiple comparison and one-way ANOVA using Tukey’s correction was used for multiple comparisons where applicable. Qualitative variables were compared with Parson’s χ2 test. Contingency analysis was performed by chi squared and Fisher’s exact test (two-sided). Based on a comparison between two groups by t-test, a sample size of: n=12/group for was shown to have >90% power to detect differences in heart weight means at P<0.05, n=7/group was shown to have >90% power to detect difference between E/A means at P<0.05; 5 mice/group was shown to have >90% power to detect difference between infarct size as well as phosphocreatine means with a significance level of P<0.05. Statistical analysis was performed using GraphPad Prism (v10.2.2) software. Differences were considered significant when P<0.05.

## Results

### Higher mortality and poorer functional outcome in STEMI type 2 diabetic patients

Forty patients (mean age 65 ± 1.3) were present in type 2 diabetes STEMI group and 40 patients without diabetes (mean age 65 ± 1.3) were in the control arm. Baseline characteristics are shown in Table 1. T2D patients were significantly obese (BMI 33 ± 0.5 kg/m^2^ vs 29 ± 0.4 kg/m^2^, P=0.0001) but were comparable to control group in terms of hypertension and plasma lipid profile (Table 1, Table 2). At hospital admission, as well as three days post-admission, T2D patients had significantly altered plasma parameters versus non-diabetic patients (Table 2, Supplementary Table 1). T2D patients had significantly elevated plasma glucose as acute hyperglycaemia is common in STEMI patients^29^.

**Table 2.** Patient plasma profile at the time of hospitalization Data are presented as mean ± SEM. Normality of data distribution was examined using Shapiro– Wilk’s normality test. Comparison between two groups was performed by Student’s t-test (Gaussian data distribution) or Mann-Whitney U test when data was nonnormally distributed. *p<0.05 vs control. Abbreviations: CRP-C reactive protein; MCV – Mean Corpuscular volume; Na – Sodium, Ca – Calcium; Mg – Magnesium; P – Phosphorus; AST – Aspartate aminotransferase; ALT – Alanine aminotransferase; HDL – High-density lipoprotein cholesterol; LDL – Low-density lipoprotein cholesterol.

|  | **Control**<br>(n=40) | **Diabetic**<br>(n=40) | P value |
| --- | --- | --- | --- |
| ***Hospital admission timepoint*** |  |  |  |
| Erythrocytes (x 10<sup>9</sup>/L) | 4.25±0.09 | 4.54±0.22 |  |
| Haemoglobin (g/L) | 129.2±3.1 | 129.1±2.4 |  |
| Haematocrit (g/L) | 0.38±0.008 | 0.38±0.007 |  |
| Thrombocytes (x 10<sup>9</sup>/L) | 286.7±19.6 | 270.1±9.9 |  |
| CRP (mg/L) | 3.6±0.5 | 4.2±0.6 |  |
| Leukocytes (x 10<sup>9</sup>/L) | 10.64±0.7 | **\*11.66±0.5** | 0.0131 |
| MCV (fL) | 89.71±1.21 | **\*87.97±0.78** | 0.0431 |
| Glucose (mmol/L) | 7.53±0.2 | **\*14.18±1.31** | 0.0001 |
| Urea (mmol/L) | 6.48±0.4 | 7.45±0.5 |  |
| Creatinine (mmol/L) | 79.43±3.5 | 93.83±6.6 |  |
| Potassium (mmol/L) | 3.75±0.08 | **\*4.01±0.09** | 0.049 |
| Na (mmol/L) | 139.7±0.4 | **\*137.5±0.6** | 0.049 |
| Ca (mmol/L) | 2.22±0.05 | 2.34±0.034 |  |
| Mg (mmol/L) | 0.81±0.01 | **\*0.74±0.02** | 0.002 |
| P (mmol/L) | 0.97±0.05 | 1.09±0.06 |  |
| AST (U/L) | 24.34±1.8 | 30.06±2.34 | 0.029 |
| ALT (U/L) | 23.50±1.89 | **\*29.49±2.16** | 0.0452 |
| Dyslipidaemia | 39 (97%) | 35 (87%) |  |
| Cholesterol (mmol/L) | 5.7±0.2 | 5.6±1.6 |  |
| Triglycerides (mmol/L) | 2.4±0.2 | 2.5±0.3 |  |
| HDL (mmol/L) | 1.1±0.05 | 1.2±0.06 |  |
| LDL (mmol/L) | 3.6±0.2 | 3.4±0.2 |  |

Coronary angiography identified multivessel disease in both patient groups, mostly 3 vessel CAD (75% in diabetics vs 57% in controls, Figure 1B) with predominant culprit lesion in right coronary artery (RCA) and left anterior descending (LAD) artery (Supplementary Table 2). Whilst both group of patients had comparable door-balloon time (Figure 1C), T2D patients had significantly higher markers of myocardial damage such as circulating troponin (Figure 1D) and plasma markers which are known to correlate with poorer ACS outcomes – increased plasma leukocytes, ALT and lower Mg (Table 2). We also found that 3 days post-STEMI, women with T2D maintain higher leukocyte count vs control group suggestive of enhanced inflammation (Supplementary Table 1).

**Figure 1.**
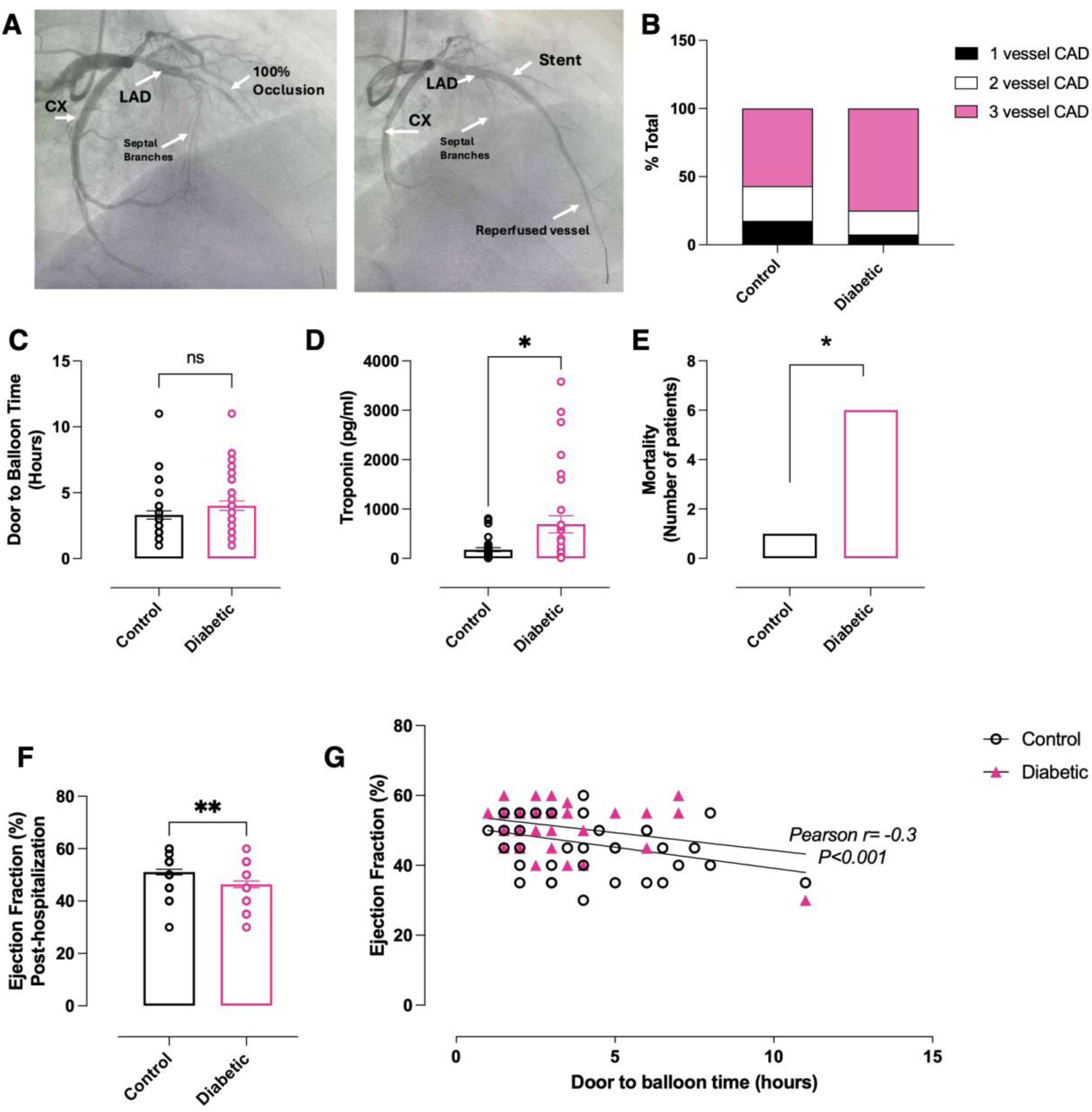
STEMI outcome in diabetic and non-diabetic female patients. **A)** Representative image of total LAD occlusion and restored coronary reperfusion post-stent placement in diabetic STEMI patient. **B)** Number of vessels affected by CAD. **C**) onset (door) to balloon time. **D**) Circulating troponin levels at the time of hospitalization. **E**) In-hospital mortality. **F**) Left ventricular ejection fraction 3 days post pPCI. **G**) Door to balloon time significantly correlates with plasma troponin. Data are presented as mean ± SEM. Normality of data distribution was examined using Shapiro–Wilk’s normality test. Comparison between two groups was performed by Student’s t-test (Gaussian data distribution), or Mann-Whitney U test when data was non-normally distributed. *p<0.05, n=40 STEMI patients with diabetes, n=40 STEMI non-diabetic controls.

T2D patients had significantly higher post-STEMI mortality vs non-diabetic control group (Figure 1E) accompanied by markedly increased akinesis of the infarcted region post pPCI (Table 3) leading to lower left ventricular ejection fraction (LVEF, Figure 1F) and increased usage of implantable cardioverter defibrillators in T2D group (Table 3). Furthermore, there was a significant negative correlation between door-balloon time vs LVEF (Figure 1G).

**Table 3.** Coronary angiography and percutaneous coronary intervention outcomes Data are presented as mean ± SEM. Normality of data distribution was examined using Shapiro– Wilk’s normality test. Comparison between two groups was performed by Student’s t-test (Gaussian data distribution) or Mann-Whitney U test when data was non-normally distributed. Contingency analysis by chi-squared and Fisher’s exact test (two-sided). *p < 0.05 vs control. Abbreviations: CABG-coronary artery bypass CRT-cardiac resynchronization therapy, PCI-percutaneous coronary intervention, ICD-Implantable cardioverter defibrillator, **PCI-additional stent requirements for non-culprit lesions and **Time to PCI number of days required to carry out additional procedures

|  | **Control**<br>**(n=40)** | **Diabetic**<br>**(n=40)** | **P value** |
| --- | --- | --- | --- |
| Total stent length (mm) | 29.3±2.2 | **\*21.6±1.6** | 0.0075 |
| Stent diameter (mm) | 3.5±0.08 | 3.3±0.09 |  |
| Total number of Stents<br>in pPCI | 38 (95%) | 36 (90%) |  |
| CABG after index<br>procedure | 1 | 3 |  |
| ICD | 0 | **\*4** | 0.0402 |
| CRT | 0 | 1 |  |
| Akinesis | 5 | **\*24** | 0.0001 |
| **Elective PCI after<br>index procedure | 14 | 17 |  |
| **Time to elective PCI<br>(days) | 22±5 | 27±5 |  |

Whilst the total number of stents as well as stent diameter was comparable between groups (Table 3), total stent length was shorter in T2D group (Table 3) as they were implanted with shorter stents to reduce the subsequent complex revascularization risk. Furthermore, T2D patients required additional elective PCI (17 vs 14 in non-diabetic group, Table 3). This is indicative of additional requirements for stents in non-culprit lesions, and presence of multivessel disease.

### Phenotypic features of female db/db mice mimic characteristics of women with T2D

20-week female db/db mice (Figure 2A) developed a distinct diabetic phenotype characterised by obesity (Figure 2B), cardiac hypertrophy (Figure 2C, 2D), hyperinsulinaemia and marked hyperglycaemia (Figure 2E, 2F). Plasma markers of renal function (urea, creatinine), liver function (ALT, AST) and free fatty acids were comparable between the groups (Supplementary Figure 1).

**Figure 2.**
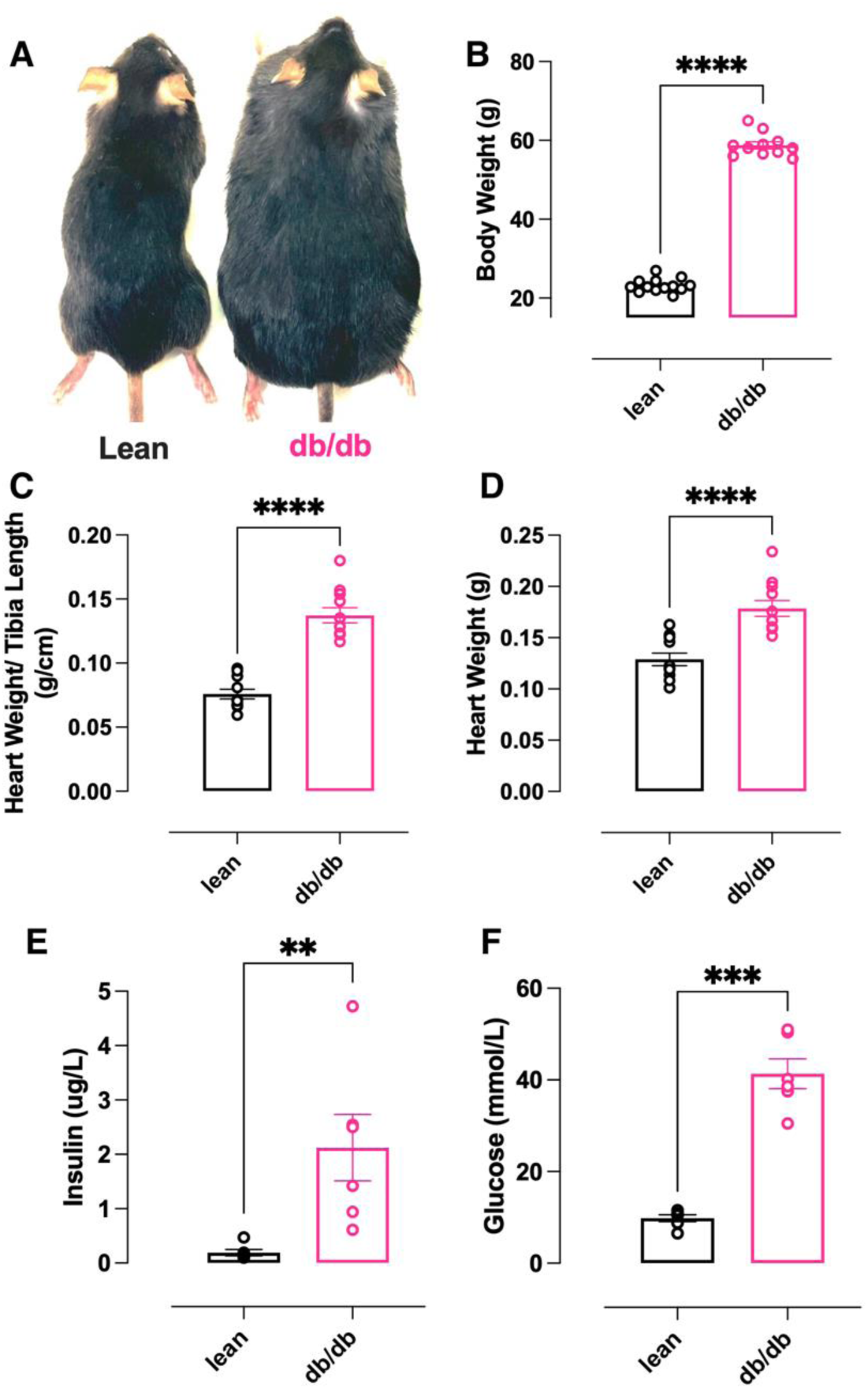
Female db/db mouse morphological features. **A**) Representative image of age-matched lean and db/db female mice. **B**) Body weight measurements, **C**) index of hypertrophy (heart weight/tibia length), and **D**) heart weight, (n=12 lean, n=12 dbdb). **E**) plasma insulin concentration and **F**) plasma glucose concentration, (n=6 lean, n=6 dbdb). Data are presented as mean ± SEM. Normality of data distribution was examined using Shapiro–Wilk’s normality test. Comparison between two groups was performed by Student’s t-test (Gaussian data distribution), unequal variance t-test, or Mann-Whitney U test when data was non-normally distributed, *p<0.05.

In addition to cardiac hypertrophy, echocardiographic assessment identified significant functional alterations (Figure 3) including reduced heart rate, increased LV internal diameter (LVID), end diastolic volume, stroke volume, mitral valve (MV) E/e’, IVRT, and MV deceleration time. However, ejection fraction was comparable to lean controls suggestive of HFpEF with diastolic dysfunction rather than HFrEF phenotype.

**Figure 3.**
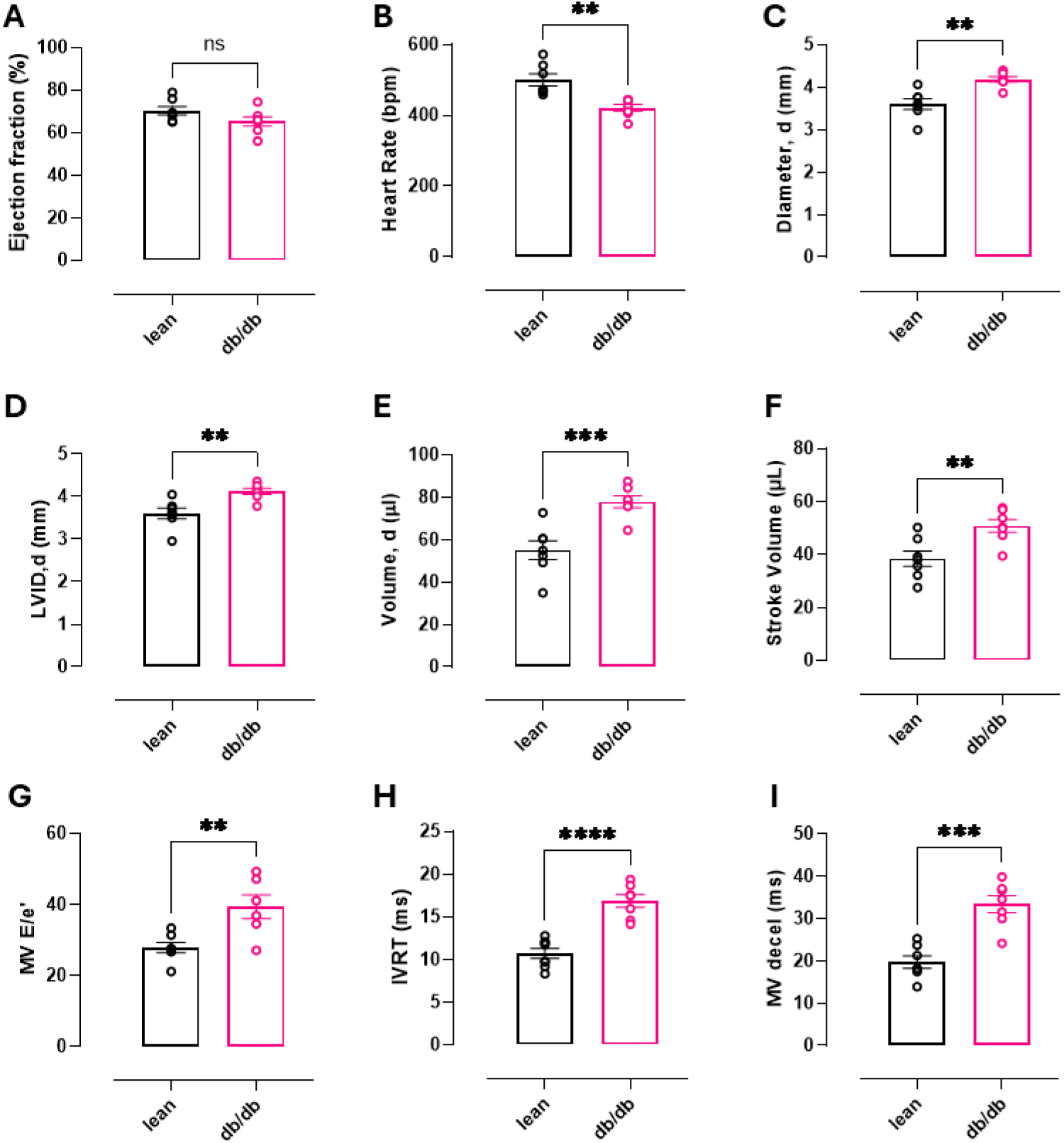
Echocardiographic assessment of in vivo cardiac function. (**A-F**) Echocardiography measurements taken from parasternal short axis view: **(A)** Ejection fraction, **(B)** Heart rate, **(C)** Heart diameter in diastole, **(D)** LV internal diameter (LVID) in diastole, **(E)** End diastolic volume and **(F)** Stroke volume. **(G-I)** Assessment of diastolic function taken from apical four chamber view: **(G)** Ratio of E mitral valve flow Doppler E wave to mitral annulus tissue Doppler e’ wave, **(H)** Isovolumetric relaxation time and **(I)** mitral valve deceleration time. Data are presented as mean ± SEM. Normality of data distribution was examined using Shapiro–Wilk’s normality test. Comparison between two groups was performed by Student’s t-test (Gaussian data distribution), unequal variance t-test, or Mann-Whitney U test when data was non-normally distributed, *p<0.05 (lean n=7, db/db n=7).

### db/db mice have altered metabolic profile and increased susceptibility to ischemia/reperfusion injury

Development and progression of T2D in db/db mice leads to significant alterations in cardiac (Figure 4A), liver and skeletal muscle metabolomic profile (Figure 4B, 4C). Myocardial metabolomic profile of db/db mice is characterised by significantly decreased NADH and creatine as well as significantly elevated succinate and branched chain amino acids (leucine, valine and isoleucine).

**Figure 4.**
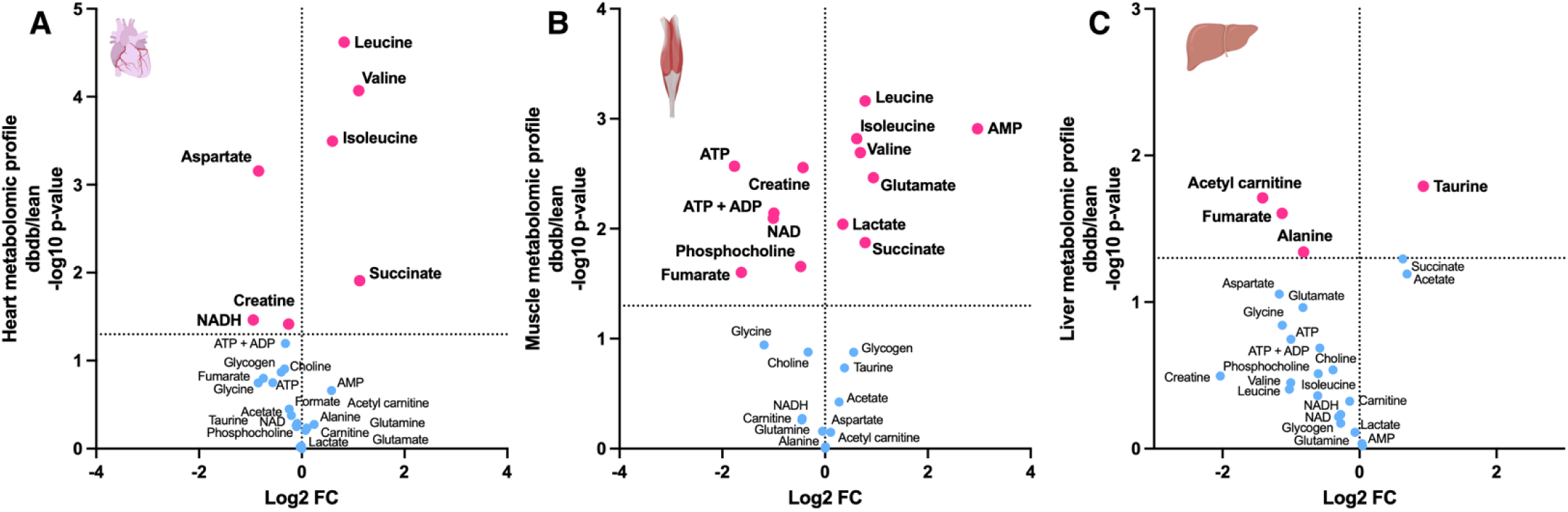
Metabolomic profile of heart, skeletal muscle and liver determined by ^1^H NMR spectroscopy Metabolomic profile of. **A)** Heart (n=5/group), **B)** Skeletal muscle (n=6/group) and **C)** Liver tissue (n=7/group) from female db/db mice compared to lean controls, measured by ^1^H NMR spectroscopy. Each point represents the fold change (FC) of a metabolite plotted against the associated level of statistical significance for the change analysed by t-test (horizontal dashed line indicates p=0.05 threshold).

Skeletal muscle metabolomic profile is severely altered with reduced energetic reserved (creatine, ATP, ADP, NAD), lipid and TCA cycle intermediates (Figure 4B). db/db skeletal muscles show marked elevation of lactate, succinate, AMP as well as amino acids (glutamate, valine, leucine, isoleucine, Figure 4B). T2D in db/db mice also caused altered liver metabolomic profile with altered levels of taurine, fumarate, acetyl carnitine and alanine (Figure 4C).

Whilst baseline *ex vivo* cardiac function of Langendorff-perfused db/db hearts was comparable to lean controls (Figure 5A), when subjected to 20-min ischemia db/db hearts show poorer functional recovery upon reperfusion (Figure 5). Alongside significantly reduced LVDP (Figure 5A), across reperfusion db/db hearts exhibit increased and irregular heart rate indicative of arrhythmogenesis (Figure 5B). TTC staining (Figure 5C) revealed that db/db hearts had significantly bigger infarct size compared to controls (Figure 5D).

**Figure 5.**
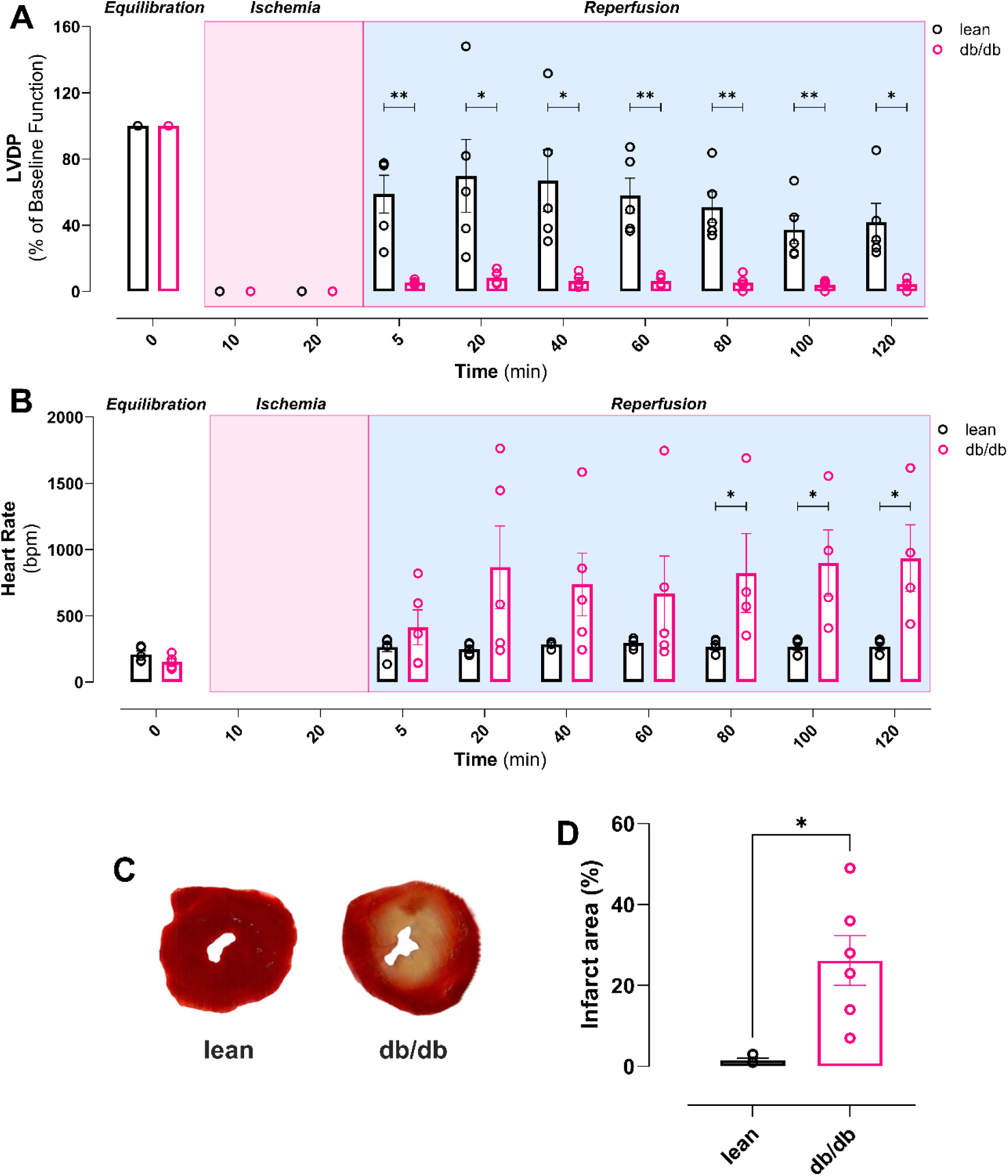
Female db/db mouse hearts have severely impaired post-ischemic recovery. **A**) Left ventricular developed pressure (LVDP) during equilibration, ischemia and reperfusion. **B**) Heart rate during equilibration, ischemia and reperfusion. **C**) Representative image of lean and db/db TTC stained hearts **D**) Infarct area quantification. Data are presented as mean ± SEM. Normality of data distribution was examined using Shapiro–Wilk’s normality test. Comparison between two groups was performed by Student’s t-test (Gaussian data distribution), unequal variance t-test or Mann-Whitney U test when data was non-normally distributed. *P<0.05, (n=5 lean, n=5 db/db).

## Discussion

Our study shows that, despite comparable time to reperfusion (DTB), diabetic female STEMI patients have poorer outcomes with higher mortality, reduced post-pPCI LVEF, increased plasma markers of cardiac damage and increased requirement for implantable cardioverter-defibrillator devices compared to non-diabetic controls (summarized in Figure 6). We also identified blood marker parameters suggestive of significantly higher systemic inflammation, cardio-hepatic syndrome as well as lower circulating magnesium, a known independent predictor of electrocardiographic no-reflow and long-term mortality in diabetic women. Acute stress of STEMI resulted in hyperglycaemia in the diabetic group despite the use of glycaemic control medications.

**Figure 6.**
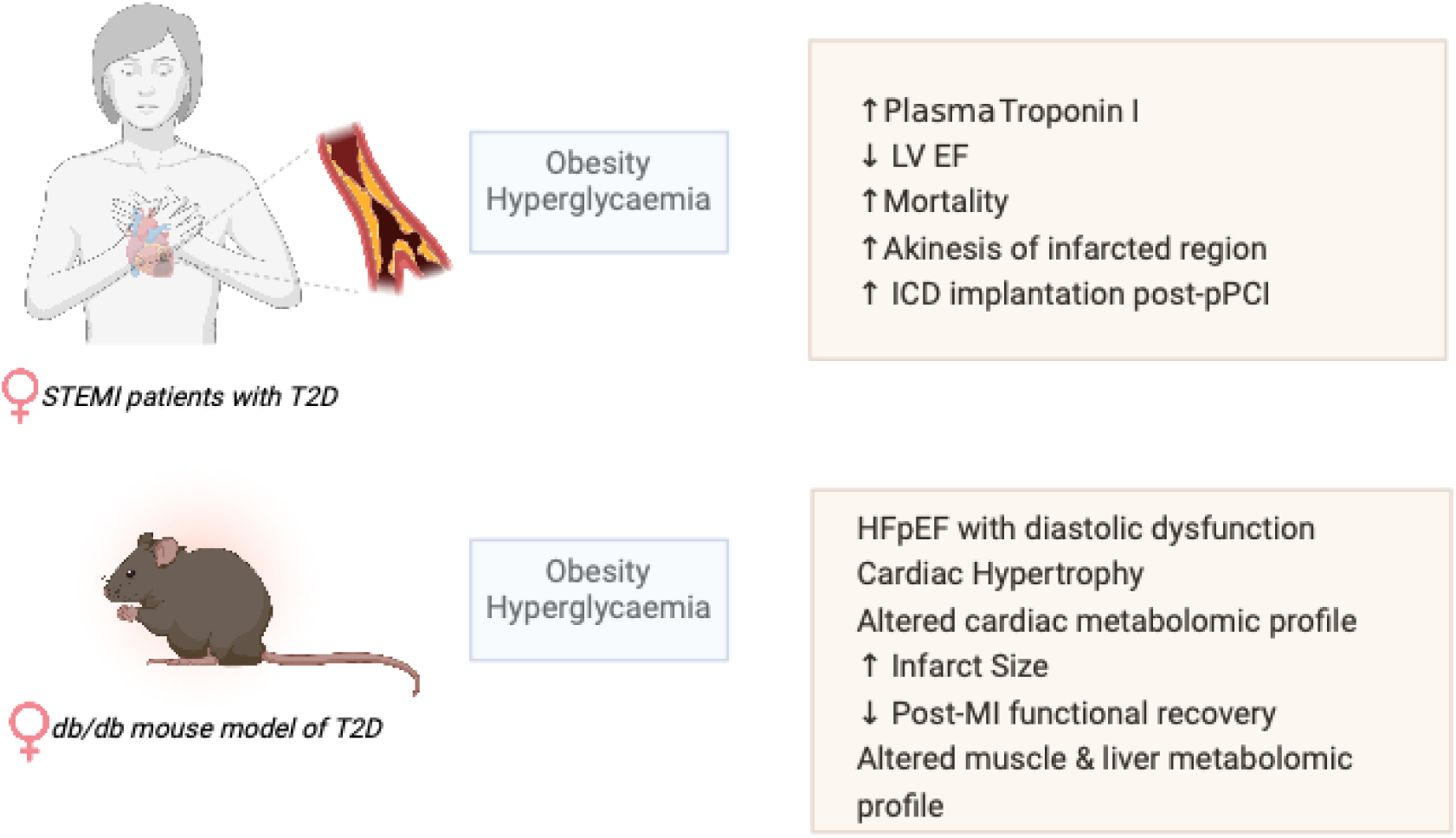
Schematic summary of post-MI changes in female patients and mice with T2D.

Comparable to diabetic female humans, female db/db mice exhibit obesity, hyperglycaemia and insulin resistance. Female db/db mice developed cardiac structural changes typical of HFpEF (myocardial hypertrophy, impaired diastolic function, LV stiffness). Furthermore, T2D led to significant perturbation of skeletal muscle, liver, and heart metabolism indicative of cardiometabolic syndrome. These metabolic comorbidities are known risk factors in the development of HFpEF. This cluster of multi-organ metabolic disturbances creates a complex network of pathways that exacerbate cardiac stress, contributing to the severe myocardial dysfunction post-I/R (poorer functional recovery and larger infarct size versus non-diabetic controls). This is the first study to combine both female T2D patients and db/db mice showing that T2D aggravates the course of myocardial infarction worsening the prognosis. Crucially, our study shows that in diabetic females longer DTB time is not the key determinant of poor STEMI recovery. Pre-clinical animal models of T2D studies seldom include female animals, leading to a gap in available robust data for the better understanding of the impact of biological sex in T2D cardiomyopathy^30^. Our study highlights female db/db mice as a feasible experimental model for the investigation of the impact of acute myocardial infarction in T2D with the outcome comparable to humans.

Coronary lesions in T2D patients typically appear long and diffuse, more likely in small-diameter arteries^31^. Consequently, lesion complexity could be relatively presented by the length and diameter of the stent. We observed no difference in diameter of stent and similar number of stents in pPCI between the groups. Furthermore, total stent length was significantly shorter in T2D patients. Given that the stent length significantly increases the thrombotic risk and the risk of stent-driven recurrent ischemic events^32^, T2D STEMI patients were implanted with shorter stents, most often “spot stenting” to reduce the subsequent complex revascularization risk. Nevertheless, we have observed that T2D patients required further PCI or CABG on additional non-culprit lesions post-pPCI vs non-diabetics.

To what extent do the observed post-MI reductions in LVEF, higher mortality, and elevated troponin levels in diabetic women reflect a direct effect of T2D versus larger myocardial areas at risk or microvascular dysfunction? Our data, showing a similar burden of three-vessel coronary artery disease in diabetic and non-diabetic patients, refutes the hypothesis that a larger myocardial area at risk is the primary determinant of the observed disparities in outcome. There is also no significant difference in terms of blood vessels affected as both groups predominantly had lesions on the RCA and LAD. The comparable distribution of culprit lesions (LAD and RCA) in both groups, considering the LAD’s supply to approximately 50% of the left ventricle, suggests no significant difference in myocardial involvement between diabetic and non-diabetic patients. Comparable revascularization strategies, including the number of stents used, stent diameter, and extent of additional procedures (PCI or CABG) on non-culprit vessels, were observed in both groups. This suggests that differences in the myocardial area at risk are not responsible for the increased incidence of reduced LVEF and mortality observed in the diabetic group. Whilst we acknowledge that diabetic HFpEF is characterised by underlying reduction in capillary density due to pericyte apoptosis, poorer endothelial dysfunction and increased oxidative stress^33^, we can exclude the potential impact of microvascular obstruction on the diabetic group STEMI outcome as myocardial infarction without obstructive CAD (MINOCA) and spontaneous coronary artery dissection (SCAD) patients were not included^34,35^.

Our study has shown that despite comparable treatment window, STEMI patients with T2D had alterations in blood marker parameters which correlate with increased cardiac damage during MI: increased ALT, increased leukocytes and lower magnesium. Hepatic injury secondary to HF is well described, however, only limited data exist about the possible impact of ACS on the liver^36^. Here we have identified possible cardio-hepatic interaction in STEMI patients with T2D as they had higher circulating level of ALT on admission. Previous work has shown that greater severity of cardiac dysfunction was associated with worse elevation of liver enzymes including ALT^36,37^. The combination of decreased LVEF and venous congestion was shown to be associated with elevated circulating liver enzymes suggesting a possible cardio-hepatic syndrome among patients with STEMI^37^. Elevated ALT was also independently associated with increased in-hospital all-cause mortality in patients with STEMI^37^. Furthermore, liver metabolomic profile of db/db mice has revealed that the development of T2D leads to impaired metabolism including depletion of lipid, amino acid and Krebs cycle intermediates as well as elevation of taurine.

Regarding the inflammatory mechanisms involved in STEMI, the leukocyte count in T2D group was significantly higher on admission. Higher leukocyte count has been shown to correlate with STEMI size and is associated with the poor prognosis and mortality^38^. Sex differences also impact the inflammatory response after STEMI. Women were found to present with STEMI with higher lymphocyte counts^39^. On hospital admission, T2D STEMI patients also had lower circulating Mg. Lower circulating magnesium on admission in STEMI patients is an independent predictor of electrocardiographic no-reflow and long-term mortality as it has been shown to have pro-thrombolytic, pro-inflammatory and pro-atherogenic effects^40^.

STEMI induces acute hyperglycaemic response, driven by cortisol and adrenaline release, which is significantly amplified in diabetic patients due to their compromised glucose control^41^. This stress-induced hyperglycaemia is associated with worse outcomes (increased mortality and morbidity). The effective management of acute hyperglycaemia is thus vital for improving outcomes in diabetic women presenting with STEMI. However, variations in clinical presentation and higher prevalence of comorbidities complicate treatment, underscoring the need for gender-specific considerations in therapeutic strategies. Persistent hyperglycaemia following STEMI poses long-term implications as well. It can hinder recovery, increase the risk of subsequent cardiovascular events, and exacerbate long-term complications associated with diabetes, making it essential to prioritise glycaemic control to improve overall prognosis^41^.

In diabetic cardiomyopathy, worse MI outcome is in part caused by the loss of metabolic flexibility, energetic deficit and mitochondrial dysfunction^42^. Cardiac metabolomic profile of db/db mice is consistent with these observations as characterized by reduced creatine content. It has previously been shown that creatine is protective and anti-apoptotic during myocardial ischemia as it enhances energy reserve and cell survival^43^. T2D caused significant alteration in myocardial amino acid metabolism. Of note, branched chain amino acids (isoleucine, leucine, valine) were markedly elevated indicative of chronically impaired regulation leading to their accumulation^44^.

db/db hearts were also characterized by elevated succinate in normoxia. It has been shown that individuals with T2D and obesity exhibit elevated levels of succinate in the circulation, urine, faeces, liver and retina^45^. We have previously shown in male db/db mice that the altered cardiac metabolism including elevated myocardial succinate is accompanied by the presence of dysregulated T-cell-mediated cardiac inflammation^46^. Given that succinate is not only oxidized but effluxed from the hearts^19^, this normoxic succinate accumulation in db/db hearts could also act as a pro-inflammatory stress signal worsening the MI outcome observed in patients and mice^47,48^. Succinate was shown to be the only metabolite significantly increased in coronary sinus blood in STEMI patients correlating with the extent of acute myocardial injury^49^. Furthermore, diabetic myocardium has been shown to be more susceptible to symptomless silent myocardial ischemia^50^ due to underlying vascular disorders which could be driving the accumulation of myocardial succinate preceding the onset of MI. Furthermore, diabetic myocardium is characterized by Na_i_ accumulation^3^. Elevated myocardial Na_i_ was recently shown to cause pseudo-hypoxia and stabilization of HIF-1α despite normal tissue oxygenation, to decrease Gibb’s free energy of ATP hydrolysis, increases the TCA cycle intermediates succinate and fumarate, decreases ETC activity at Complexes I, II and III, and causes a redox shift of CoQ to CoQH ^51^. Thus, myocardial Na_i_ elevation could be responsible for the succinate accumulation in female db/db hearts.

Diabetic HFpEF is increasingly recognized as a multiorgan disorder rather than solely a cardiac dysfunction^52^. Given the metabolic crosstalk between organs, and to better understand mechanistic links between systemic metabolic dysfunction and cardiac performance, we investigated the metabolic signature of liver and skeletal muscles in female db/db mice. Skeletal muscle was found to have the most severe and extensive perturbation in metabolomic profile. Exercise intolerance in HFpEF is often linked to skeletal muscle dysfunction rather than cardiac output limitations alone. T2D was shown to lead to greater skeletal muscle atrophy and metabolic dysfunction which may be causing poorer exercise tolerance in HFpEF^53^. Decreased glycolysis and increased β-oxidation were previously shown to lead to increased branched-chain amino acid concentrations in T2D skeletal muscles^54^.

T2D women with STEMI often face disparities in treatment they receive. They may receive less aggressive therapy compared to men leading to worse outcomes^15^. This reflects a disadvantage of women with T2D based on behavioural, treatment or societal aspects^15^. Diabetic women may delay seeking care due to social responsibilities, such as caregiving roles, or due to underestimating the severity of their symptoms. However, our study has shown comparable door-balloon time between T2D and non-diabetic patient groups. Women with T2D have higher prevalence of atypical STEMI symptoms (fatigue, nausea, shortness of breath) rather than a classic chest pain thus delaying diagnosis and timely intervention^8,15^. These atypical symptoms are mostly secondary to peripheral diabetic neuropathy, which can, due to an increase oxidative stress, reduce perception of pain, nerve growth factors and mitochondrial dysfunction^55,56^.

This study is limited by its single-centre design, which may restrict the generalisability of our findings to other healthcare settings or diverse populations. Although our patient cohort is adequately sized to identify significant differences in outcomes, a larger, multi-centre cohort could yield more definitive conclusions. Additionally, our analysis did not include male patients or mice, preventing us from making any conclusions about potential inter-sex differences.

## Conclusions

Considering the rising prevalence of T2D and its associated cardiovascular risk, the impact of T2D on STEMI in women is of critical importance for public health. Understanding the interplay between T2D and STEMI in women can lead to better prevention strategies, earlier recognition of symptoms and more effective treatment options. Future research focusing on sex-specific strategies in T2D management is essential for improving cardiovascular disease outcomes in women.

## Declarations

### Ethics approval and consent to participate

The study protocol was approved by the Ethics Committee of University Clinical Centre Tuzla (Ref. 02-09/2-166-2/24) and all methods were performed in accordance with their guidelines and regulations. All animal experiments were in keeping with the UK Home Office Animals (Scientific Procedures), 1986.

## Consent for publication

All authors consent the manuscript submission and publication.

## Availability of data and materials

All data deposited open access on dryad.com DOI: 10.5061/dryad.3n5tb2rts.

## Authors’ contributions

I.I, M.Y, E.C. F.C, H.A.P, D.A. performed the experiments. I.I. M.Y. E.C. D.A processed and analysed the data. I.I.M.Y.D.A, HAP, MPM. wrote the main manuscript text. DA, MY prepared all the figures. All authors reviewed the manuscript.

## Data Availability

All data produced in the present study are available upon reasonable request to the authors

## List of abbreviations

T2D: type 2 diabetes
CAD: coronary artery disease
ACS: acute coronary syndrome
MI: myocardial infarction
HF: heart failure
STEMI: ST segment elevation myocardial infarction
pPCI: primary percutaneous coronary intervention
I/R: ischemia reperfusion
LVEF: left ventricular ejection fraction
LV: left ventricle
KH: Krebs–Henseleit
LVDP: left ventricle developed pressure
TTC: triphenyltetrazolium chloride
NMR: nuclear magnetic resonance
SEM: standard error of mean
ANOVA: analysis of variance
RCA: right coronary artery
LAD: left anterior descending
ALT: alanine aminotransferase
AST: aspartate aminotransferase
LVID: left ventricle internal diameter
MV: mitral valve
HFpEF: heart failure with preserved ejection fraction
HFrEF: heart failure with reserved ejection fraction

## Acknowledgements

We would like to thank Dr Nasima Kanwal and School of Physical and Chemical Sciences NMR Facility, Queen Mary University of London.

