## Supplementary material for "Type 2 diabetes worsens the outcome of ischemia/reperfusion in female STEMI patients and female db/db mice with HFpEF cardiometabolic phenotype": Online Supplement

| | **Control** (n=40) | **Diabetic** (n=40) | P value |
|---|---|---|---|
| ***3 days post hospital admission*** | | | |
| Erythrocytes (x $10^9$/L) | 4.20±0.07 | 4.24±0.08 | |
| Haemoglobin (g/L) | 128.9±2.44 | 125.5±1.89 | |
| Haematocrit (g/L) | 0.38±0.006 | 0.37±0.005 | |
| Thrombocytes (x$10^9$/L) | 253.2±14.8 | 247.8±12.4 | |
| Leukocytes (x $10^9$/L) | 8.79±0.52 | ***10.01±0.49** | 0.0079 |
| MCV(fL) | 88.55±1.91 | 88.58±0.81 | |
| Glucose (mmol/L) | 6.04±0.18 | ***11.49±0.68** | 0.0001 |
| Urea (mmol/L) | 6.29±0.40 | 7.63±0.59 | |
| Creatinine (mmol/L) | 74.03±2.39 | ***93.79±7.47** | 0.018 |
| Potassium (mmol/L) | 4.16±0.08 | 4.36±0.08 | |
| Na (mmol/L) | 138.5±0.71 | 138.7±0.69 | |
| Ca (mmol/L) | 2.26±0.02 | 2.32±0.035 | |
| Mg (mmol/L) | 0.84±0.01 | 0.81±0.02 | |
| P (mmol/L) | 1.26±0.025 | ***1.12±0.06** | 0.0103 |
| AST (U/L) | 86.84±8.42 | ***63.31±6.42** | 0.0292 |
| ALT (U/L) | 42.36±3.54 | 38.50±3.01 | |

**Supplementary Table 1. Patient plasma profile 3 days post-hospitalization**
Data are presented as mean ± SEM. Normality of data distribution was examined using Shapiro–Wilk's normality test. Comparison between two groups was performed by Student's t-test (Gaussian data distribution) or Mann-Whitney U test when data was non-normally distributed.

| **Culprit vessel** (number of patients) | **Control** (n=40) | **Diabetic** (n=40) |
|---|---|---|
| LM | 1 | 0 |
| LAD | 14 | 16 |
| Prox LAD | 7 | 9 |
| Mid LAD | 6 | 8 |
| Dist LAD | 0 | 0 |
| CX | 3 | 5 |
| Prox CX | 0 | 3 |
| Dist CX | 3 | 1 |
| OM | 1 | 2 |
| RCA | 22 | 17 |
| Prox RCA | 6 | 10 |
| Mid RCA | 14 | 5 |
| Dist RCA | 2 | 2 |

**Supplementary Table 2**. Number of culprit vessels affected by occlusions
LAD – left anterior descending, CX - circumflex, OM – obtuse marginal, RCA – right coronary artery, Prox-proximal, Mid-medial, Dist-distal

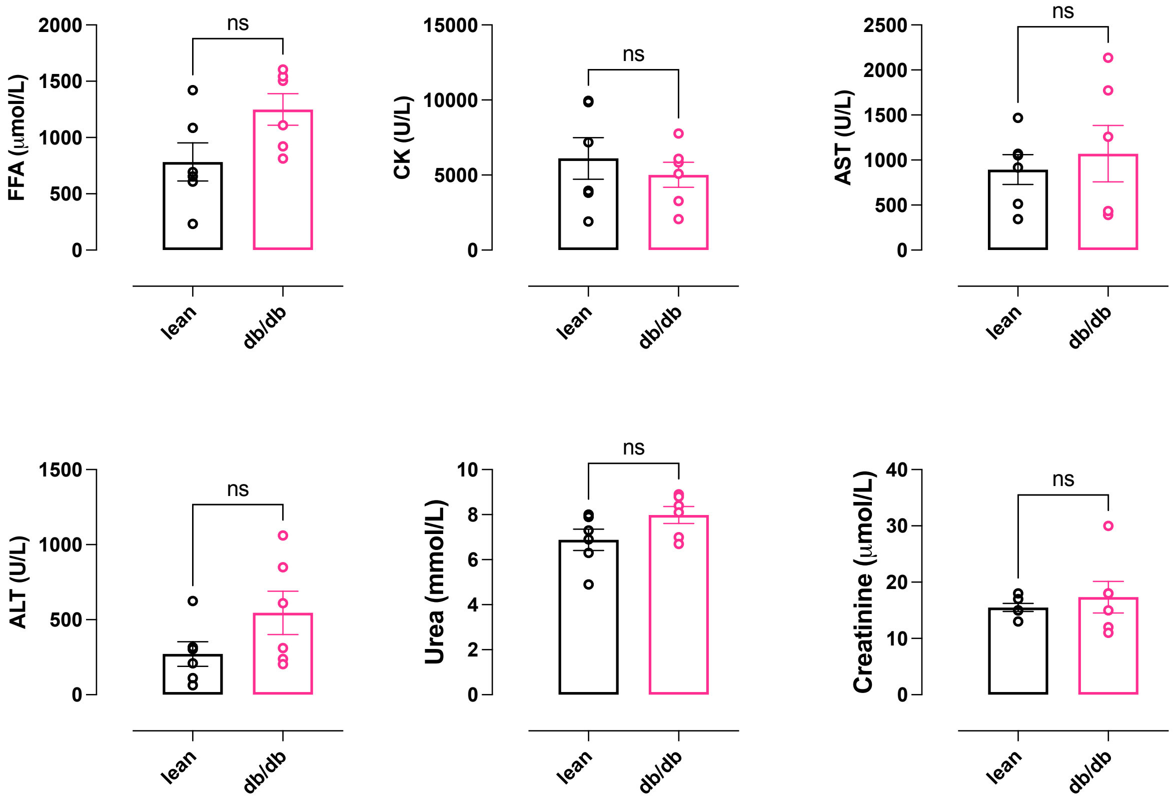


**Supplementary Figure 1.** Mouse plasma profile lean vs db/db 20 week female mice. (n=6 lean, n=6 dbdb). Data are presented as mean ± SEM. Normality of data distribution was examined using Shapiro–Wilk's normality test. Comparison between two groups was performed by Student's t-test (Gaussian data distribution), unequal variance t-test, or Mann-Whitney U test when data was non-normally distributed, *p<0.05.
